# Health workforce preparedness for sepsis care in Bahrain: a cross-sectional survey of guideline knowledge, attitudes, and practices

**DOI:** 10.64898/2026.09.10.26362792

**Authors:** Khadija Aziz Oameer Ebrahim, Ammar Ali Jaber, Rafiullah Baig Mirza

**Author notes:** Correspondence: Rafiullah Baig Mirza,.

## Abstract

Sepsis and septic shock remain critical medical emergencies that require timely and effective interventions. This study evaluates the knowledge, attitudes, and practices (KAP) of healthcare professionals in Bahrain concerning the management of sepsis and septic shock. A cross-sectional survey was conducted among a sample of 260 healthcare professionals who consented to participate out of 310 approached, yielding a response rate of 83.9%. Data were collected using a structured, self-administered questionnaire designed to ensure anonymity and consistency in responses. Data were analyzed using descriptive and inferential statistics via IBM SPSS version 29. Most participants demonstrated sound general knowledge of sepsis, with 90.7% correctly identifying broad-spectrum antibiotics as the first-line treatment. However, awareness of certain clinical tools was lower—only 44.6% recognized the qSOFA score and 60% understood the use of capillary refill time. Knowledge levels were significantly higher among males (77.4%) than females (70.6%, p = 0.043), and among those with advanced degrees (p < 0.001), especially doctorate holders (84.5%). Physicians had the highest average knowledge scores (91.0%), followed by PharmD holders (82.2%). Positive attitudes toward early sepsis recognition were reported by 82.2% of participants, particularly among those with doctoral education and over 21 years of experience. In practice, 72.7% routinely assessed for organ dysfunction, and 51.9% initiated antibiotic therapy within the first hour of diagnosis. The study identified significant gaps in diagnostic tool familiarity and clinical practice consistency. These findings underscore the importance of initiatives to enhance adherence to sepsis management protocols in Bahrain.

## Introduction

Sepsis is a life-threatening organ dysfunction in response to an infection, and it remains a leading cause of morbidity and mortality worldwide [1]. Sepsis remains a critical global health challenge with substantial morbidity and mortality worldwide. In 2017, an estimated 48.9 million incident cases of sepsis were recorded globally, resulting in 11.0 million sepsis-related deaths, representing 19.7% of all global deaths [2]. The World Health Organization reports that sepsis affected 49 million people globally in 2017, with approximately 20% of annual global deaths attributable to this condition [3]. Age-standardized sepsis incidence has demonstrated a declining trend, falling by 37.0% from 1990 to 2017, while mortality decreased by 52.8% during the same period [2]. Although sepsis is a major burden in Low- and Middle-Income Countries (LMIC), high income countries also face difficulties in early recognition and management [2]. Although the Kingdom of Bahrain has a well-developed health system, delays to sepsis diagnosis, variable adherence to evidence-based guidelines, and limited resources still hinder its management [4]. Adequate understanding of knowledge, attitudes, and practices (KAP) of healthcare professionals (HCPs) in Bahrain regarding sepsis will ensure better patient outcomes. Evidence based sepsis guidelines have been provided by Surviving Sepsis Campaign (SSC) since 2004, SSC versions that follow include new research and best practice recommendations [5]. According to the 2021 SSC guidelines, early recognition, prompt administration of antibiotics, and brief cooperative force management are to prevent mortality [5]. Although this is not optimal in the world, a large part of that is due to lack of adequate training and awareness or systemic limitations [6]. Like other countries, the translation of these recommendations into clinical practice is a challenge, and therefore evaluation of KAP towards these recommendations among the healthcare workforce of Bahrain is very important to identify gaps for development of targeted interventions [7].

Sepsis and septic shock are major factors that cause both morbidity and mortality in patients in Bahrain, especially those in ICU. Analysing the situation in the King Hamad University Hospital, Shirazy et al., [8] identified that sepsis and septic shock were also a significant load to ICU, the mortality rates being from 20% to 40%, depending on the disease severity and the time span of the treatment received. Studies on healthcare-associated infections in Bahrain’s hospitals reveal that various practices in the clinical settings include hand hygiene and infection measures but the early detection of sepsis and prescribed antibiotics are not practiced uniformly. For instance, a study conducted in three Saudi hospitals highlighted on the absence of some hospital antibiotic policies and this may influence delays on the right management of sepsis [9]. There have also been other researches concerning low compliance to pneumonia and sepsis care in some hospitals in the gulf countries including Bahrain where the mean stays and mortality was also reported to be higher [10].

Research has been conducted on sepsis and septic shock management in Bahrain. For instance, Shirazy et al., [8] conducted a prospective study that showed Hypernatremia can be an independent predictor of poor outcome in septic and septic shock patients in the ICU in King Hamad University Hospital, Bahrain. A few studies have revealed that healthcare professionals in Bahrain generally agree on the need for more sepsis training [11,12,13]. Notwithstanding, knowledge gaps exist regarding the annual sepsis mortality rate and specific management protocols, particularly regarding fluid resuscitation and antibiotic administration in Bahrain. Alsalman et al., [14] also noted that there is limited data on the epidemiology, clinical manifestations and outcomes of patients with invasive aspergillosis, a serious fungal infection, can lead to sepsis, in Bahrain. No study has attempted to relate knowledge and attitudes of healthcare professionals in Bahrain toward sepsis management to their education level, profession, and gender. Moreover, there is limited information on the extent to which healthcare professionals adhere to best practices in sepsis management in Bahrain as well as barriers that hinder their adherence. Resolving this research gaps will help improve sepsis and septic shock management in Bahrain. This is imperative as sepsis and septic shock are among the most prevalent causes of intensive care unit (ICU) admissions, accounting for approximately 10-50% of the mortality rate [15].

Therefore, the objectives of this cross-sectional study are to investigate the influences of education level, profession, and gender on knowledge and attitudes of healthcare professionals toward sepsis management, to determine the extent to which healthcare professionals adhere to best practices in sepsis management in Bahrain, and to determine the barriers that hinders the adherence of healthcare professionals to sepsis management guidelines in Bahrain.

## Materials and Methods

### Research design

This research employed a cross-sectional study design to assess the knowledge, attitudes, and practices (KAP) of healthcare professionals regarding the management of sepsis and septic shock, in alignment with recent standard protocols. The cross-sectional design enabled data collection from multiple healthcare facilities at a single point in time, providing a quantitative measure of the KAP levels of healthcare workers (HCWs) in Bahrain. This approach was suitable for identifying knowledge or practice gaps without requiring follow-up and allowed for a rapid evaluation of existing sepsis care processes and identification of areas where educational or clinical improvements might be necessary [16]. The study was conducted over a six-month period, allowing sufficient time for participant recruitment, questionnaire dissemination, and data collection across different shifts and departments.

### Data collection tool

A structured, self-administered questionnaire was developed for this study to assess healthcare professionals’ knowledge, attitudes, and practices (KAP) regarding sepsis and septic shock management. Item content was derived from the Surviving Sepsis Campaign guidelines and current clinical recommendations. The questionnaire comprised 45 items across three domains: 18 knowledge items with true/false/I don’t know response options; 5 attitude items rated on a five-point Likert scale from Strongly Disagree to Strongly Agree; and 22 practice items rated on a five-point frequency scale from Never to Always. The domains covered sepsis definitions, early recognition, scoring tools, guideline-based interventions, professional confidence, institutional support, fluid resuscitation, antibiotic timing, and diagnostic monitoring.

### Questionnaire development and validation

Questionnaire validation was undertaken through a sequential, multistage process. First, an independent panel of experts in critical care, infectious diseases, sepsis management, clinical pharmacy, nursing, and questionnaire methodology evaluated each item for relevance, clarity, and representativeness using a four-point rating scale. The item-level content validity index (I-CVI) was calculated as the proportion of experts who assigned a rating of 3 or 4. Items with an I-CVI of at least 0.78 were retained, whereas items below this threshold were revised or removed following panel discussion. The scale-level content validity index using the average method (S-CVI/Ave) was also calculated, with a value of at least 0.90 considered satisfactory. Expert comments and the resulting item revisions were documented.

Face validity and response-process validity were subsequently examined through cognitive interviews with healthcare professionals who were representative of the intended respondents. Participants were asked to explain their interpretation of each item, the reasoning underlying their selected response, and their understanding of the response options. Ambiguous wording, unfamiliar terminology, double-barrelled questions, and response options that did not adequately reflect clinical practice were revised. Cognitive interviewing was conducted iteratively until no substantial interpretation problems were identified.

The revised questionnaire was then pilot-tested among healthcare professionals who were excluded from the main survey. Pilot testing assessed questionnaire completion time, missing responses, item comprehension, response distributions, and potential floor and ceiling effects. Feedback from the pilot participants was used to simplify wording, clarify clinical terminology, improve response options, and refine the questionnaire before its administration to the main study sample.

Internal consistency was evaluated separately for each questionnaire domain. The Kuder–Richardson 20 coefficient was used for the dichotomously scored knowledge items, while McDonald’s omega and Cronbach’s alpha were calculated for the attitude and practice domains. Coefficients between 0.70 and 0.95 were considered acceptable. Corrected item–total correlations were also examined, and items with values below 0.30 underwent clinical and statistical review. Because the knowledge domain covered several distinct components of sepsis management, its internal-consistency coefficient was interpreted cautiously and was not used as the sole criterion for removing clinically important items.

Temporal stability was evaluated by readministering the unchanged questionnaire to a subsample of participants approximately two weeks after the initial administration. Intraclass correlation coefficients based on absolute agreement were calculated for the domain scores, with values of at least 0.75 considered evidence of acceptable test–retest reliability. Weighted kappa coefficients were also calculated for individual ordinal items where appropriate.

The dimensional structure of the attitude and practice domains was investigated using exploratory factor analysis based on polychoric correlations. Sampling adequacy was assessed using the Kaiser–Meyer–Olkin statistic and Bartlett’s test of sphericity. The number of factors retained was determined through parallel analysis and inspection of the scree plot. Because correlations among the underlying dimensions were anticipated, an oblique rotation method was applied. Items with primary factor loadings below 0.40, substantial cross-loadings, or weak conceptual relevance were reviewed and revised or removed as appropriate.

The factor structure identified through exploratory factor analysis was subsequently evaluated using confirmatory factor analysis. Model fit was assessed using the comparative fit index, Tucker–Lewis index, root mean square error of approximation, and standardized root mean square residual. CFI and TLI values of at least 0.90 and RMSEA and SRMR values of 0.08 or lower were considered indicative of acceptable model fit. Where the sample size permitted, measurement invariance across professional groups was also examined. Following completion of these analyses, poorly performing items were revised or removed, and the reliability of the finalized questionnaire domains was reassessed.

### Study area

Data collection took place primarily at MKCC, with fewer than 10% of participants recruited from another private hospital in Bahrain, which requested to remain unnamed. The individuals who participated in the pilot study were excluded and then the survey link was shared with participants from both medical centres. Facilities to be included in the study were chosen based on the presence of steering protocols for sepsis care, preferably based sepsis protocols such as the SSC. Specifically, this criterion was important in the determination of compliance to benchmark protocols in sepsis care. In this way, the study enrolled hospitals where the workforce is assumed to have exposure to sepsis management guidelines, either through adopting or advocating for best practices in sepsis care.

### Study population and subject criteria

The study target population included all healthcare personnel in the care of sepsis and septic shock patients. This included physicians, nurses and all other health care personnel involved in patient care most especially in the intensive care unit and emergency units. Including only the professionals who are directly involved in sepsis treatment, the study aimed to get data from the respondents with practical experience in handling sepsis patients.

### Inclusion criteria

Participants were eligible for the study if they met the following criteria: direct involvement in sepsis management; minimum of one year of clinical experience; formal training or certification in sepsis management, and informed consent.

### Exclusion criteria

The following criteria were used to exclude certain individuals from the study: non-clinical roles; students and interns, and decline of consent

### Sample size calculation

The sample size for this study was calculated to ensure adequate power to detect statistically significant differences in KAP scores among healthcare professionals.

The formula used for estimating sample size in cross-sectional studies was applied as follows:

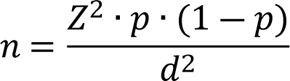

Where: Z = 1.96 (corresponding to 95% confidence level), p = 0.5 (assumed prevalence of adequate knowledge, to maximize variability), and d = 0.0575 (margin of error, 5.75%) Therefore, a minimum sample size of 290 healthcare professionals was required. This target sample ensured broad representativeness across professional categories and healthcare settings, enabling subgroup analyses by role (physician, nurse, pharmacist, clinical pharmacist) and sector (public vs. private). While two healthcare institutions in Bahrain were included, the actual number of responses obtained was 260, representing 89.66% of the calculated target sample. Despite falling slightly short of the estimated target, the achieved sample size provided sufficient diversity and analytical robustness for descriptive and inferential analyses. Non-response bias was minimized through informed consent, explanation of study purpose, and assurances of anonymity and confidentiality.

### Sampling method and subject recruitment

A stratified random sampling method was employed to ensure representativeness across key professional groups involved in sepsis and septic shock management. The target population was stratified into four subgroups: physicians, nurses, pharmacists, and clinical pharmacists. This stratification allowed for adequate subgroup analysis, while reducing selection bias and enhancing the external validity of the study. After stratification, random sampling was applied within each professional group to select participants from the two participating healthcare facilities in Bahrain. Quotas were proportionally allocated to reflect the distribution of staff across these roles. Recruitment was facilitated in collaboration with hospital administrations, department heads, and professional group coordinators. The research team contacted potential participants through official hospital emails, bulletin notices, and direct communication with departmental leaders. All potential respondents received a study information sheet describing the objectives, significance, and procedures of the study, along with assurances of confidentiality and voluntary participation. Digital informed consent was obtained prior to questionnaire access. Participants were also reminded that their identities would remain anonymous and that they were free to withdraw at any time.

### Data collection procedures

Data collection was conducted using the finalized, self-administered questionnaire hosted on a secure Google Forms platform. Access was restricted to invited participants to preserve response integrity and anonymity, and the survey link was distributed by email through department heads and hospital administrative contacts in accordance with the sampling framework. The 25 healthcare professionals who completed the pilot validation were not eligible for the main survey. Participants required approximately 10-15 minutes to complete the questionnaire. Data collection remained open for six weeks to include different work shifts and clinical rotations, and periodic reminders were sent to improve participation. Technical assistance was available when required.

## Data analysis

All quantitative data were analyzed using IBM SPSS Statistics (Version 29). Prior to inferential testing, all continuous variables were examined for distributional assumptions using the Shapiro-Wilk test to determine normality.

The analysis proceeded across several key domains of the KAP framework:

1. Descriptive Statistics: Frequencies, percentages, means, and standard deviations were calculated to summarize the demographic characteristics of participants and their responses to individual items within the knowledge, attitude, and practice sections.
2. Knowledge Scoring: Each correct answer in the knowledge section was assigned one point. A total knowledge score was generated for each respondent. Descriptive analysis was followed by one-way ANOVA to compare knowledge scores across demographic subgroups (e.g., professional role, years of experience, sepsis training). Post-hoc comparisons were conducted when significant differences were identified.
3. Attitude Scoring: Responses to Likert scale items were numerically coded (1 = Strongly Disagree to 5 = Strongly Agree). Individual scores were averaged to calculate an overall attitude score for each participant.
4. Practice Analysis: Frequencies and percentages were computed for practice items. Responses were also evaluated against core components of the Surviving Sepsis Campaign (SSC) guidelines to assess compliance levels. Where relevant, cross tabulations were performed to examine associations between practice behaviors and professional background or prior training.
5. Inferential Statistics: One-way ANOVA was applied to compare KAP scores across key demographic groups, where appropriate, post-hoc Tukey’s tests were used to identify specific group differences. Statistical significance was determined at a p-value < 0.05.

### Ethical considerations

Ethical approval was obtained from [FULL NAME OF ETHICS COMMITTEE] (approval number: [REQUIRED]) before data collection. All participants provided digital informed consent before accessing the questionnaire. The study was conducted in accordance with the Declaration of Helsinki.

## Results

### Demographic characteristics

The study enrolled 260 healthcare professionals with a cross-sectional survey on the knowledge, attitudes, and practices on sepsis and septic shock management based on recent guidelines. The demographic profile of the participants was examined to bring out a background to the study results. As presented in Table 2, out of all the respondents, 66% (172) identified as female and 34% (88) as male. A predominance of females was noted based on respondents’ gender since a majority of health care employees; especially the nurses formed the biggest profession in this research. The largest number of respondents, 48.8% (n=126), self-identified as Asian, which can be regarded as the most significant racial category of the study. The second largest group that participated in the study was the Middle Eastern/North African with 38.4 % (n=99) of the total participants. White/Caucasian participants accounted for 5.8% (n=15). The rest of the participants were black/African American, Hispanic/Latino, native American/Alaskan, or native Hawaiian/pacific islanders and these represented a small proportion of the overall racial representation.

Of all respondents, most of them, 38.8 % (n=101) stated that they had a bachelor’s degree. The second largest category was postgraduate students having a post graduate degree of master’s degree constituting 32.2% (83) of the total subjects. Of all the participants, 15.9% (n = 41) had completed their education at the Diploma level. Using the above definitions, it was also noted that 12.8% (n=33) participants completed a Doctorate level of education.

All the target participants were selected from different healthcare disciplines that contributed to the care of patients with sepsis and septic shock. The largest group among the respondents was comprised of the nurses who represented 59.1%, n = 154. Doctors that participated in the research represented 19.5 % of the respondents, n = 51. Specifically, 16% (n=42) of the participants were pharmacists; 5% (n=13) of them held a Doctor of Pharmacy (PharmD) degree. As for the workplace, 90.3% of the respondents work in specialized hospitals and 9.7% in general hospitals. Also, the vast majority of the participants, 95.8% were from a public hospital while only 4.2% from a private hospital. The survey of respondents in terms of experience revealed that 20.4% (n=53) of them had professional experience of less than two years. Of the participants, 23.1% (n=60) said that they had between two and five years of experience. Participants with professional working experience of between six and ten years formed 20.8% (54 respondents) of the sample. Among the participants, 29.2% (n=76) had between 11-20 years of experience while 4.2% (n=11) had 21 years and above of overall healthcare practice.

### Knowledge of sepsis and septic shock management

The survey of the knowledge among healthcare professionals on sepsis and septic shock management involved 18 questions. The participants’ responses to the statements are provided in Table 1 with the number of participants selecting “True”, “False”, and “I Don’t Know”. The results are organized under four major aspects of sepsis: Knowledge about Sepsis and Septic Shock, Diagnostic Methods, Primary Care of Sepsis, and Advanced Management of Sepsis. The breakdown of response distribution is given in Table 1 below.

**Table 1.**
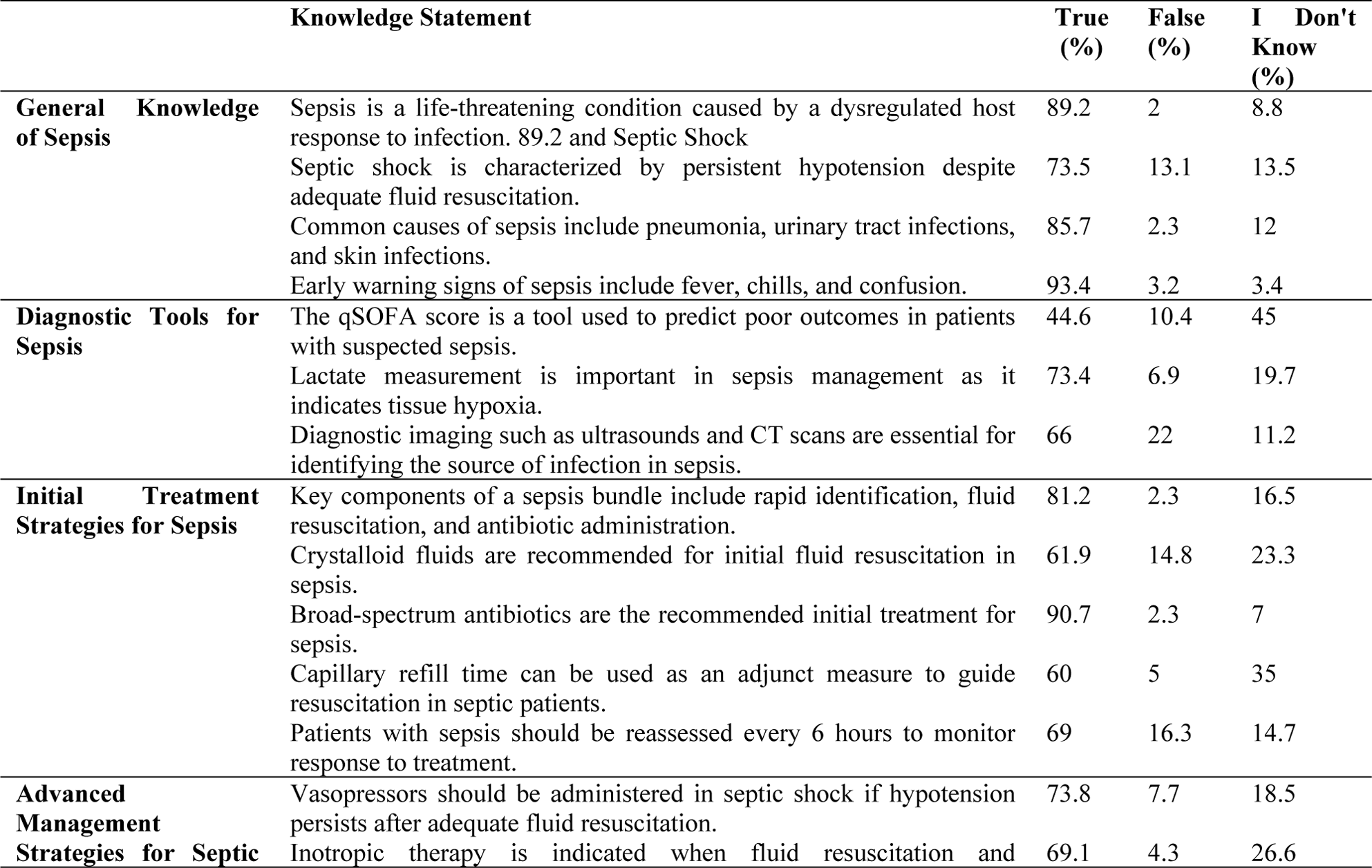
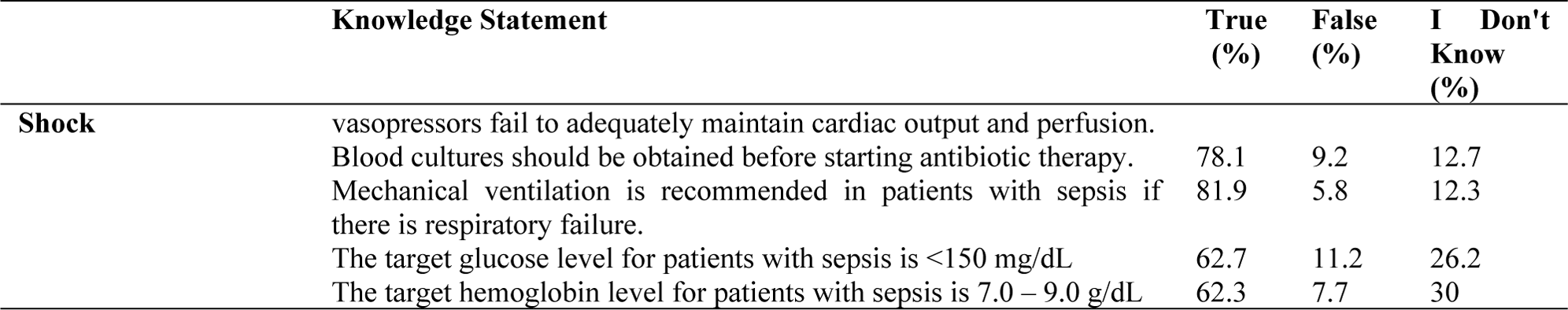
Knowledge of healthcare professional participating in KAP survey for Sepsis and Septic Shock Management (n=260)

There was an overall satisfactory performance of the participants on basic knowledge about sepsis and how it should be identified. Most (89.2%) of the respondents agreed that sepsis is a lethal condition that arises from the host’s immune response to an infection. Likewise, 73.5% of the participants correctly defined septic shock as hypotension that does not respond to fluids for more than 2 hours, while 13.5% of them were unsure. Common infections that cause sepsis included pneumonia which was known by 85.7% of the respondents. This indicates the participants’ rather detailed knowledge of the clinical picture of sepsis, which is essential for timely and accurate diagnosis.

Awareness of diagnostic markers and scoring systems of sepsis was low. 73.4% of the participants were right about lactate measurement as being the indication of tissue hypoxia but 19.7% of the participants had no clue about lactate measurement in sepsis. There was a fairly poor knowledge of the qSOFA score where 44.6% of the participants affirmed it as a tool that assesses prognosis in septic patients with organ dysfunctions and 45% of the participants were unsure. This could be an educational issue since qSOFA is actually recommended for risk stratification when it comes to sepsis sight. In regard to diagnostic imaging, 66.8% appreciated the significance of modality such as ultrasound and CT scan for the identification of sepsis source while 22% gave wrong answer about significance of imaging in sepsis related workup.

The participants had good knowledge about basic principles of sepsis management since 81.2% of them were aware that the components of sepsis management include prompt identification, fluids, and antibiotics. Also, 90.7% of the participants identified that broad-spectrum antibiotics were the drugs of choice for treating any infection. But only 61.9% could accurately identify crystalloid fluids as the appropriate choice and 23.3% of the candidates were equivocal about the fluids to be used for resuscitation. The awareness of Capillary Refill Time (CRT) as an adjunct measure for resuscitation was comparatively low with 60% of the participants recognizing its use while 35% were unsure indicating a possibility of knowledge deficit with regards to assessing bedside perfusion.

The understanding about use of vasopressors and inotropic therapy was moderate: while 73.8% realized that vasopressor is appropriate in septic shock when hypotension persists despite fluid therapy, only 18.5% were unsure. Also, 69.1% correctly identified inotropes when initial fluid and vasopressors prove ineffective while only 26.6% remained ambivalent. There was poor knowledge on reassessment in sepsis management whereby 69% of respondents knew that patients with sepsis should undergo reassessment every six hours and 16.3% gave an incorrect statement. Regarding the blood cultures, only 78.1% of participants knew that blood culture samples should be collected prior to administering antibiotics in the management of sepsis while 12.7% were ambiguous.

Moreover, mechanical ventilation was recognized by healthcare providers with high percentage of 81.9% agreeing with the necessity of using it in respiratory failure confirming excellent understanding of ventilatory support in septic critically ill patients. The participants’ awareness of the target physiological parameters was dispersed. As for the management of the target glucose concentration under 150 mg/dL, only 62.7% of respondents gave the correct answer, and 26.2% expressed uncertainty. Similarly, 62.3% correctly noted the target haemoglobin range (7.0-9.0 g/dL), although 30% were not certain about it; this highlights that regarding metabolic and oxygenation targets in sepsis management, learners are still in deficit.

### Knowledge statistical analysis

This section presents the results on the level of knowledge regarding sepsis and septic shock among participants, categorized by gender, educational attainment, professional role, and years of experience. Knowledge scores were classified as Low (<60%), Moderate (60 – 79.99%), and High (≥80%) based on raw mean percentage scores.

As shown in Table 2, male participants (n = 88) had a mean knowledge score of 77.39% (SD = 24.58), while female participants (n = 171) scored 70.57% (SD = 27.34). Both groups were classified under Moderate Knowledge. The difference in scores between males and females was statistically significant (p = 0.043), though no post-hoc analysis was applicable.

**Table 2.**
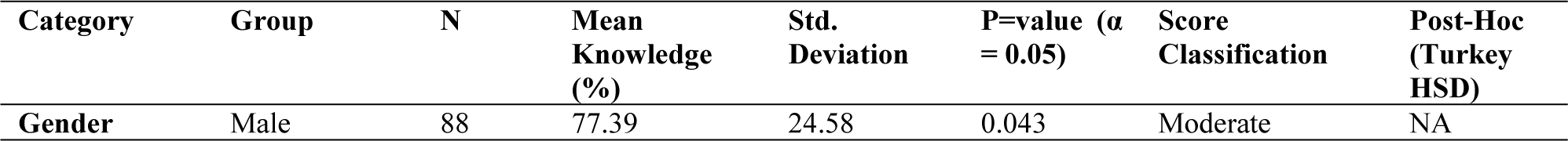
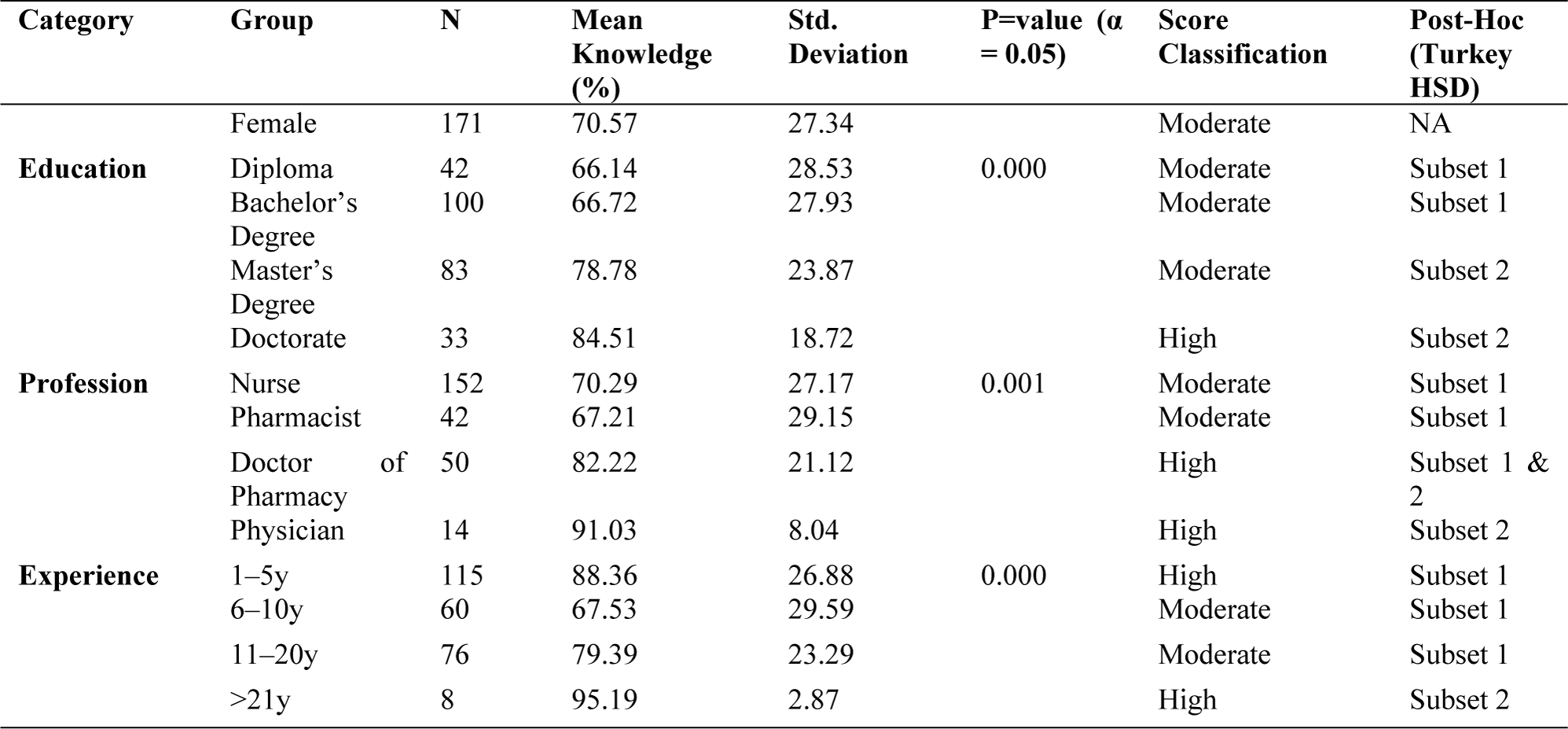
Summary of mean knowledge scores and ANOVA between different groups and t-test for Gender category (n=260)

Participants with a Doctorate (n = 33) scored 84.51% (SD = 18.72), falling under the High Knowledge category. Those with a master’s degree (n = 83) scored 78.78% (SD = 23.87), classified as Moderate Knowledge. Bachelor’s degree holders (n = 100) and Diploma holders (n = 42) scored 66.72% (SD = 27.93) and 66.14% (SD = 28.53), respectively, both within the Moderate Knowledge category. The differences were statistically significant (p < 0.001), with post-hoc grouping placing Diploma and Bachelor’s in Subset 1, and Master’s and Doctorate in Subset 2.

The highest mean knowledge score was observed among Physicians (n = 14) with 91.03% (SD = 8.04), followed by Doctor of Pharmacy holders (n = 50) with 82.22% (SD = 21.12). Both were classified under High Knowledge. Nurses (n = 152) scored 70.29% (SD = 27.17) and Pharmacists (n = 42) scored 67.21% (SD = 29.15); both groups were classified under Moderate Knowledge. Group differences were statistically significant (p = 0.001). Post-hoc comparisons grouped Nurses and Pharmacists in Subset 1, Physicians in Subset 2, and Doctor of Pharmacy across Subsets 1 and 2.

Participants with >21 years of experience (n = 8) scored the highest at 95.19% (SD = 2.87), and those with 1–5 years (n = 115) scored 88.36% (SD = 26.88). Both were classified under High Knowledge. Participants with 11–20 years (n = 76) scored 79.39% (SD = 23.29), falling within the Moderate Knowledge range, as did those with 6–10 years (n = 60) who scored 67.53% (SD = 29.59). These differences were statistically significant (p < 0.001). Post-hoc analysis grouped the 1–20-year categories into Subset 1, and those with >21 years in Subset 2. Participants with more than 21 years of experience demonstrated the highest mean knowledge score (95.19%), supporting the assumption that prolonged clinical exposure enhances familiarity with complex conditions such as sepsis. This finding aligns with expectations that senior clinicians possess deeper insights into diagnosis and treatment pathways due to accumulated experience.

### Attitudes toward sepsis and septic shock management

The attitude was measured using five statements in terms of participants’ views towards the value of early recognition and treatment of sepsis, level of confidence to identify sepsis, clarity of guidelines, institutional support, and peers support for sepsis, as shown in Table 3. All the options were given on a Likert scale of 1-5, where 1 represents ‘strongly disagree’ and 5 which represents strongly agree’. Majority of the respondents (82.2%) either agreed or strongly agreed with early recognition and treatment of sepsis to enhance the patients’ prognosis. Only a small portion of the respondents – 7.8% – strongly disagreed with the statement and 10% of respondents remained neutral, which shows the majority of the respondents acknowledged sepsis as an urgent condition that requires immediate attention.

**Table 3.** Summary of mean attitude scores and ANOVA between different groups and t test for Gender category (n=260)

| Category | Group | N | Mean Attitude (%) | Std. Deviation | P=value ($\alpha = 0.05$) | Attitude Classification | Post-Hoc (Turkey HSD) |
| --- | --- | --- | --- | --- | --- | --- | --- |
| **Gender** | Male | 88 | 75.03 | 25.78 | 0.518 | Positive | NA |
|  | Female | 171 | 77.53 | 14.99 |  | Positive | NA |
| **Education** | Diploma | 42 | 77.43 | 16.52 | 0.001 | Positive | Subset 1 & 2 |
|  | Bachelor's Degree | 100 | 71.52 | 21.62 |  | Positive | Subset 1 |
|  | Master's Degree | 83 | 79.71 | 18.27 |  | Positive | Subset 1 & 2 |
|  | Doctorate | 33 | 84.85 | 12.72 |  | Positive | Subset 2 |
| **Profession** | Nurse | 152 | 74.87 | 20.18 | 0.009 | Positive | Subset 1 |
|  | Pharmacist | 42 | 77.95 | 13.03 |  | Positive | Subset 1 |
|  | Doctor of Pharmacy | 50 | 81.12 | 18.68 |  | Positive | Subset 1 & 2 |
|  | Physician | 14 | 90.77 | 7.55 |  | Positive | Subset 2 |
| **Experience** | 1–5y | 115 | 76.27 | 15.58 | 0.026 | Positive | Subset 1 |
|  | 6–10y | 60 | 75.47 | 17.81 |  | Positive | Subset 1 |
|  | 11–20y | 76 | 76.05 | 24.72 |  | Positive | Subset 1 |
|  | >21y | 8 | 91.47 | 13.26 |  | Positive | Subset 2 |

The level of confidence in recognizing the risks of sepsis according to the participants’ responses was not uniform. A majority of them at 66.5% had confidence as indicated by agree/strongly agree while 22.7% were neutral, and 10.8% had no confidence as indicated by disagree/strongly disagree. This can imply that while most healthcare providers may be confident in assessing who is at risk, a significant number of healthcare providers may sometimes have doubts about it, which may indicate areas of preclinical education or training that may not have been effective. Respondents’ views on working with the recent sepsis guidelines were divided, particularly regarding their clarity and ease of implementation. While a majority (65.3 %) of the respondents described them as clear operational guidelines that can be easily applied within their organizations, 25.1% were neutral about the guidelines, and only 9.6% strongly disagreed with or disagreed on statements describing the guidelines as unclear. Whereas one quarter remains neutral, this has implications towards the degree of awareness of the guidelines or factors inhibiting implementation within clinical practice.

About the training received in the institution on how to manage sepsis and the available resources, 62.5% of the respondents agreed or strongly agreed with the statement that adequate support was provided by their institution while 23.9% gave a neutral response and 13.6% disagreed. Thus, the study shows that although most of the participants do not perceive they lack institutional support, the few who gave less than optimal ratings implied that training and resources for support should be enhanced.

Participants were also asked questions relating to support from their team when tending to sepsis cases. A larger percentage of the respondents (63.8%) stated that they had gotten adequate support from colleagues and supervisors, 26.9% said that otherwise. Only 9.3% of them disagreed or strongly disagreed, thus highlighting that, though teamwork and supervisory assistance appear to be accessible, some participants may encounter challenges to effective sepsis teamwork.

### Statistical Comparison of Attitude Scores Across Demographic Groups

Statistical analysis of participants’ attitudes towards sepsis and septic shock based on demographic and professional variables was conducted. Across all groups, the overall classification of attitude was Positive, as all mean scores exceeded the 60% threshold. However, there were statistically significant differences among groups based on education, profession, and experience.

Male participants (n = 88) had a mean attitude score of 75.03% (SD = 25.78), while female participants (n = 171) had a mean score of 77.53% (SD = 14.99). Both groups demonstrated a Positive Attitude towards sepsis care. The difference in attitude scores was not statistically significant (p = 0.518), indicating that gender did not influence participants’ attitudes.

A statistically significant relationship was found between education level and attitude (p = 0.001). Participants with a Doctorate degree (n = 33) showed the highest mean attitude score (84.85%, SD = 12.72), followed by those with a master’s degree (n = 83) at 79.71% (SD = 18.27), and Diploma holders (n = 42) at 77.43% (SD = 16.52). Those with a bachelor’s degree (n = 100) had the lowest score at 71.52% (SD = 21.62). All participants in this category had a Positive Attitude, but post-hoc analysis distinguished bachelor’s degree holders as Subset 1, Doctorate holders as Subset 2, and diploma/master’s as part of both subsets.

There was a significant difference in attitude across professions (p = 0.009). Physicians (n = 13) had the highest mean attitude score at 90.77% (SD = 7.55), followed by Doctor of Pharmacy holders (n = 50) at 81.12% (SD = 18.68), Pharmacists (n = 42) at 77.95% (SD = 13.03), and Nurses (n = 152) at 74.87% (SD = 20.18). Despite this variation, all professions maintained a Positive Attitude. Post-hoc grouping placed Nurses and Pharmacists in Subset 1, Physicians in Subset 2, and Doctor of Pharmacy holders in both subsets, reflecting overlapping score distributions.

Experience in the field also revealed statistically significant differences in attitude (p = 0.026). Participants with more than 21 years of experience (n = 15) reported the highest mean score of 91.47% (SD = 13.26), significantly exceeding other experience groups. Those with 1–5 years (n = 115), 6–10 years (n = 54), and 11–20 years (n = 76) had mean scores of 76.27% (SD = 15.58), 75.47% (SD = 17.81), and 76.05% (SD = 24.72) respectively. All groups exhibited a Positive Attitude, with post-hoc analysis separating the >21-year group into Subset 2 and the rest into Subset 1.

### Practice in sepsis and septic shock management

The survey results (Table 4) demonstrate healthcare professional practices in sepsis and septic shock management appear in Table 4. The frequency distribution of management practices used by respondents appears in Table 4 starting from never to always.

**Table 4.** Practice in Sepsis and Septic Shock Management.

| Question | Never (%) | Rarely (%) | Sometimes (%) | Often (%) | Always (%) |
| --- | --- | --- | --- | --- | --- |
| Do you routinely use a sepsis screening tool (e.g., qSOFA)? | 14.3 | 11.6 | 25.1 | 23.9 | 25.1 |
| Do you regularly assess for signs of organ dysfunction in patients with sepsis? | 9.3 | 5.2 | 12.8 | 26.8 | 45.9 |
| Do you initiate fluid resuscitation within 1 hour of sepsis recognition? | 8.6 | 7.8 | 18.3 | 23.7 | 41.6 |
| Do you administer antibiotics within 1 hour of sepsis recognition? | 5.0 | 5.1 | 15.5 | 22.5 | 51.9 |
| Do you routinely obtain more than one blood culture when sepsis is suspected? | 4.3 | 5.5 | 11.0 | 27.8 | 51.4 |
| Do you actively investigate and identify the source of infection? | 6.5 | 5.0 | 6.7 | 12.4 | 69.4 |
| Do you initiate appropriate source control measures within 24 hours? | 4.6 | 10.5 | 13.1 | 19.7 | 52.1 |
| Do you monitor vital signs, lactate levels, and organ function in patients with sepsis? | 6.7 | 4.2 | 7.3 | 15.1 | 66.7 |
| Do you adjust treatment based on the patient's response to therapy? | 2.3 | 8.5 | 8.9 | 10.5 | 69.8 |
| Do you consider using corticosteroids in the management of septic shock? | 6.5 | 11.0 | 29.8 | 19.0 | 33.7 |
| Do you use ultrasound to assess fluid responsiveness in sepsis patients? | 8.9 | 9.7 | 27.9 | 20.2 | 33.3 |
| Do you routinely monitor coagulation parameters in sepsis patients? | 5.0 | 7.6 | 30.2 | 14.7 | 42.6 |
| Do you administer blood products to maintain target haemoglobin in sepsis? | 6.1 | 6.7 | 28.6 | 23.9 | 34.7 |
| Do you monitor and manage glucose levels in sepsis patients? | 8.5 | 4.5 | 8.5 | 13.5 | 65.0 |

A substantial portion of healthcare professionals makes frequent usage of sepsis screening instruments such as the qSOFA score because they assess at risk patients through these tools either constantly (25.1%) or frequently (23.9%). A substantial number of 14.3% healthcare workers report they do not utilize screening tools at any time. Among the respondents 72.7% conduct regular organ dysfunction evaluations by monitoring vital signs and signs of organ failure particularly assessing lactate levels and organ function while 45.9% perform these evaluations always. Early sepsis detection and tracking receive important emphasis through this assessment practice.

Most healthcare professionals expedite critical interventions by adhering closely to established guidelines when performing fluid resuscitation together with antibiotic administration. A substantial number of 41.6% healthcare providers prompt fluid resuscitation for sepsis patients as soon as they identify the condition within one hour while 51.9% similarly act quickly to start antibiotic treatment. The essential character of these medical practices ensures better patient results, and many professionals fulfill these protocols correctly. A minority group comprising 8.6% and 5% neglect to maintain these time-dependent treatment methods.

The investigation of infection sources together with blood culture collection shows high adherence statistics among the respondents. A majority of 51.4% of participants obtains several blood cultures right away when sepsis suspicion arises and 69.4% checks the infection source for all cases. Source control implementation within 24 hours proved vital to reducing mortality rates since 52.1% of respondents applied this measure appropriately.

The practice of adjusting treatment via patient responses is frequently used by healthcare professionals as demonstrated by the 69.8% who always modify therapy according to clinical feedback. Regular assessment of lactate levels, organ function and various vital parameters is performed by 66.7% of participants thus emphasizing continuous evaluation in sepsis management.

Several clinical practices do not appear to be regularly maintained based on this data. Ultrasound for measuring fluid responsiveness is identified as standard practice by 33.3% of medical professionals with corticosteroids being used regularly by 33.7% for septic shock therapy. The practice of coagulation parameter assessment and blood products administration stands as less widespread because only 42.6% and 34.7% of respondents conduct them routinely. The control of glucose levels demonstrates inconsistent adherence because 65% of professionals keep track of glucose levels consistently in septic patients.

### Statistical comparison of practice scores across demographic groups

The distribution of practice scores related to sepsis and septic shock management among healthcare professionals were analyzed (Table 5). Based on the classification criteria, all participant groups demonstrated a Positive Practice, as their mean scores exceeded the 60% threshold. Nevertheless, statistically significant differences were identified across gender, education, profession, and experience categories.

**Table 5.** Summary of mean practice scores and ANOVA between different groups and t test for Gender category (n=260)

| Category | Group | N | Mean Attitude (%) | Std. Deviation | P=value ($\alpha = 0.05$) | Attitude Classification | Post-Hoc (Turkey HSD) |
| --- | --- | --- | --- | --- | --- | --- | --- |
| **Gender** | Male | 88 | 83.09 | 13.52 | 0.01 | Positive | NA |
|  | Female | 171 | 76.04 | 22.8 |  | Positive | NA |
| **Education** | Diploma | 42 | 76.12 | 18.65 | 0.000 | Positive | Subset 1 & 2 |
|  | Bachelor's Degree | 100 | 71.42 | 26.07 |  | Positive | Subset 1 |
|  | Master's Degree | 83 | 85.56 | 13.24 |  | Positive | Subset 3 |
|  | Doctorate | 33 | 84.59 | 13.57 |  | Positive | Subset 2 & 3 |
| **Profession** | Nurse | 152 | 77.65 | 20.92 | 0.026 | Positive | Subset 1 |
|  | Pharmacist | 41 | 74.84 | 23.72 |  | Positive | Subset 1 |
|  | Doctor of Pharmacy | 50 | 82.54 | 14.52 |  | Positive | Subset 1 & 2 |
|  | Physician | 15 | 91.75 | 5.59 |  | Positive | Subset 2 |
| **Experience** | 1–5y | 115 | 75.41 | 22.6 | 0.008 | Positive | Subset 1 |
|  | 6–10y | 54 | 75.32 | 24 |  | Positive | Subset 1 |
|  | 11–20y | 76 | 84.08 | 12.09 |  | Positive | Subset 1 |
|  | >21y | 15 | 86.29 | 15.15 |  | Positive | Subset 1 |

The analysis revealed that male participants (n = 88) had a significantly higher mean practice score of 83.09% (SD = 13.52) compared to their female counterparts (n = 171), who scored 76.04% (SD = 22.80). The difference was statistically significant (p = 0.01), indicating that gender may influence the application of sepsis management practices. Despite this difference, both groups were classified as having a Positive Practice.

Educational level had a strong association with sepsis-related practice (p < 0.001). Participants with a master’s degree (n = 83) recorded the highest mean score (85.56%, SD = 13.24), followed closely by Doctorate holders (n = 33) with 84.59% (SD = 13.57). Diploma holders (76.12%, SD = 18.65) and bachelor’s degree holders (71.42%, SD = 26.07) had comparatively lower scores. All groups demonstrated Positive Practice, although post-hoc analysis revealed three distinct subsets, with bachelor’s degree holders forming Subset 1, master’s degree holders forming Subset 3, and Doctorate and Diploma holders overlapping both.

Professional role also exhibited a statistically significant relationship with practice scores (p = 0.026). Physicians (n = 15) reported the highest mean practice score at 91.75% (SD = 5.59), followed by Doctor of Pharmacy holders (n = 50) with 82.54% (SD = 14.52). Nurses (n = 152) and Pharmacists (n = 41) had lower scores at 77.65% (SD = 20.92) and 74.84% (SD = 23.72), respectively. Despite these variations, all professional groups maintained a Positive Practice classification. Post-hoc analysis placed Nurses and Pharmacists in Subset 1, Physicians in Subset 2, and Doctor of Pharmacy holders overlapping both.

Participants’ years of experience also had a statistically significant impact on practice scores (p = 0.008). The highest score was observed in those with more than 21 years of experience (n = 15) at 86.29% (SD = 15.15), followed by the 11–20 years group (n = 76) with 84.08% (SD = 12.09). Participants with 1–5 years (n = 115) and 6–10 years (n = 54) reported lower mean scores of 75.41% (SD = 22.6) and 75.32% (SD = 24.00), respectively. All experience groups demonstrated Positive Practice, with post-hoc analysis grouping all into Subset 1, indicating relatively consistent practices despite varied experience lengths.

## Discussion

This study reveals essential understanding about how healthcare workers approach sepsis and septic shock management. The participants showed strong grasp of basic sepsis principles, yet they displayed important gaps in their knowledge and different professional approaches and inconsistent real-world sepsis care. These study results prove that healthcare institutions must prioritize educational programs along with resource support for improvements in sepsis treatments and evidence-based guideline implementation.

Although the participants had a good understanding of the overall principles, sepsis identification, and antibiotic administration, they had poor awareness of diagnostic tools including qSOFA and lactate, assessment methods such as capillary refill time and reassessment intervals and newer management protocols involving fluids and vasopressor, inotropes, and metabolic goals. From these results, the areas of training and reiteration of lessons for enhancement may be useful.

### Knowledge of sepsis and septic shock management

The knowledge assessment for healthcare staff reveals that they have satisfactory knowledge about the basic sepsis definition, diagnosis procedural identification, and antibiotic guidelines. However, the existing questionnaire and identification knowledge gaps are extended in subsequent questions to incorporate the essentially used sepsis screening tools like qSOFA score and lactate measurement for nurses. The challenge that is likely to be experienced when using the sepsis screening tools is due to variations in nursing curricula with minimum focus on sepsis identification [17]. Prior research shows that many nurses perform poorly on the distinguishing of early and late sepsis which endorses wrong classification and treatment delays [18]. Also, the use of ‘septicemia’ as a broad term rather the specific and precise sepsis definitions hinder the clarity and treatment in clinical practice [19]. Sepsis is also a concern for pharmacists due to their low awareness of pathophysiology and protocol implementation. A systematic review of pharmacists in the management of antimicrobial stewardship suggested poor awareness of sepsis-associated infections and suboptimal approaches in prescribing antibiotics within the context of emergency care [20]. Community pharmacists with little exposure to critical care settings may not have the necessary background in sepsis guideline implementation which translates into delayed medication alterations and overall ineffective interventions by pharmacists [21]. To mitigate these gaps, there should be focused sepsis educational interventions for pharmacists as well as progression of their roles within the interprofessional teams [22]. The outcomes of the assessment in knowledge were statistically significant with educational attainment and medical rank and years of clinical practice. This study also presents medical professionals and Doctor of Pharmacy holders as groups with a better understanding of sepsis because of their higher levels of education. The results show a preference of longer practicing HC providers by ten years and over getting a better understanding of sepsis, which inflicts credit to some clinical exposure interface that leads to the growth of familiarity with knowledge on protocols [23]. Graduate programs should also focus on diploma and bachelor’s degree to fill existing skill gaps of various professionals. Community pharmacists, unlike their hospital-based or PharmD counterparts, are generally not involved in acute decision-making or direct sepsis care. Their primary roles focus on medication dispensing, patient counselling, and minor ailment management. A lack of structured clinical exposure and limited involvement in diagnostic protocols restricts their contribution to early sepsis identification or treatment decisions. Nonetheless, with proper referral pathways and continuing education, community pharmacists could potentially assist in raising early alarms or advising on symptom escalation in outpatient settings [21,22].

### Attitudes towards sepsis management

Majority of healthcare professionals had positive perceptions about sepsis including the need for quick identification and treatment for the condition. One notable limitation of the study is the potential for online response bias, as participants may have looked up answers prior to responding to the questionnaire—an inherent risk in digital data collection. One limitation highlights the practitioners’ self-confidence to identify such patients and the knowledge of sepsis protocol. A significant 65% of the participants were unsure of how they would identify sepsis since they did not receive regular training and only encounter few cases of sepsis in clinical practice [17]. Multiple studies indicated that there is an increase of confidence and competency in the proper implementation of the sepsis protocol among the trained professionals when a structured sepsis training program is provided.

The survey also revealed a significant correlation between academic progress and the perception of sepsis – healthcare providers in the doctoral level had a higher rating in terms of the belief in effective sepsis management, thus highlighting the effectiveness of education towards bearing the institutional approach to treatment. The gender differences were also found to reach significance level where the female nursing and medical staff possessed more positive attitude. With reference to the investigations made, it may be understood that female healthcare professionals are more likely to follow clinical guidelines or protocols that are in place probably because of the nursing training focus on protocol-based practice. However, even in such cases, two important issues are more or less contentious, which include institutional support and resources. According to the survey, 24% of the participants had a neutral attitude as to the credibility of the training that the institutions provided and only 13.6% disagreed with regards to the allocation of resource provisions. There is knowledge to support those leaders on different levels of the hospital need to ensure that educational initiatives are maintained, and resources are provided where needed because institution factors can limit the implementation of sepsis protocols.

### Practices of sepsis and septic shock management

Despite the certain measures of essential sepsis treatment protocols, the standard practice was not followed clinically, systematically and uniformly by many healthcare professionals. Antibiotic prescriptions (51,9% received within the 1st hour targets) and the administration of fluids (41,6% within the recommended time span/SSC) meet the standards [24]. Nonetheless, 9% of participants reported that they were never able to start with fluid-deficit replacement in the stipulated time the need for improvement. This led to nurses’ worse performance in sepsis screening tools because of the workload, self confidence, and individual differences in decision-making [18]. These scoring systems such as qSOFA also need clinical assessment, and this may not be very accurate during emergencies hence leading to variability in its use [25]. Additionally, efforts in the form of institutional requirements on providing ratio of nurses to patient can also enhance the effectiveness of screening tool and patient care outcomes as noted by Newsome et al., [26].

Still, the roles of pharmacists in sepsis care are limited; they have restricted decision-making powers, and they are not included in sepsis teams often enough. One of the critical care pharmacy services provides treatment, and a study showed that most of the pharmacists were not involved in early sepsis identification hence limiting their role in early sepsis management [21]. However, the focus of most antimicrobial stewardship initiatives is not on the sepsis alone, and the management of antibiotics is not necessarily optimized for the septic patient population in general [20]. The observation concerning the correlation between the implementation of evidence-based care practice and the educational level as well as the years of experience of the healthcare providers was highly significant. Senior doctors in training in sepsis protocol knowledge did better than the junior doctors. Despite the fact that scores increased with years of experience, there was a statistically significant difference in the scores of male and female healthcare providers: with the score of male providers being 85.7% while the score of female providers was 80% (p = 0.01). Further research should substantiate whether specific training approaches, protocols enacted by different institutions, and methods for incorporating sepsis care into clinical workflow affect the efficiency of sepsis management among various disciplines involved in sepsis care. Although physicians comprised 5.7% (n = 15) of the sample, the survey did not capture their perceptions of clinical pharmacists’ roles. This represents an important gap, given that physician endorsement is critical for integrating pharmacists into sepsis response teams. Literature suggests that physician trust and interprofessional alignment significantly enhance the implementation of pharmacist-led antimicrobial stewardship [27].

### Comparative professional roles and collaborative practice in sepsis management

Healthcare professionals demonstrate significant variations in sepsis knowledge, attitudes, and practices across disciplines. Physicians consistently exhibit superior knowledge compared to nurses and pharmacists, with emergency physicians achieving 28.4% correct identification of Sepsis-3 definitions versus 5.9% for nurses [28]. Similarly, physicians demonstrated 91.3% sufficient knowledge levels compared to 71.5% for nurses and 63.0% for pharmacists in antimicrobial management [29]. Regarding attitudes, physicians show more positive perspectives toward sepsis management than nurses [30]. A study of 200 physicians and pharmacists revealed mean attitude scores of 45.88 ± 4.00, with attitudes directly influencing practices through path coefficients of 0.91 for physicians and 0.87 for pharmacists [31]]. Recent training significantly correlates with improved knowledge across all professions [28]. Practice variations are substantial, with only 26.42% of physicians correctly identifying qSOFA score components versus minimal nurse competency [32]. Critical care nurses demonstrated moderate sepsis knowledge scores (10.56/15) but poor overall practice implementation [33]. Pharmacists show proactive practices in antimicrobial stewardship but require enhanced sepsis-specific training [31]. Nurses serve as frontline caregivers with pivotal responsibilities in sepsis recognition and management [34,35]. They function as essential members of multidisciplinary sepsis teams, often co-leading hospital sepsis programs alongside physicians [36]. Nurses play fundamental roles in detecting physiological changes that indicate sepsis onset through continuous patient monitoring and implementation of screening protocols [37]. Their responsibilities include prompt identification of sepsis through vital sign monitoring, activation of sepsis care protocols, and administration of time-critical interventions including antibiotics and fluid resuscitation [38]. Nurse-led sepsis protocols have demonstrated significant impact in reducing mortality, ICU length of stay, and improving bundle compliance [39, 40]. Pharmacists contribute significantly to antimicrobial stewardship and optimal medication management in sepsis care. They serve as drug information resources, expedite medication verification processes, and provide dosing recommendations for antimicrobial therapy [41]. Clinical pharmacists on multidisciplinary sepsis teams have been shown to decrease time to antibiotic administration, improve appropriate antibiotic selection, and reduce mortality rates [42]. Their involvement includes facilitating prompt antimicrobial delivery and supporting vasopressor management in severe cases [41]. Physicians provide clinical expertise in sepsis diagnosis, treatment decisions, and coordination of multidisciplinary care. They collaborate with nursing staff in sepsis program leadership and serve as champions across hospital departments [36]. Physicians are responsible for clinical evaluation, diagnostic workup, and implementation of evidence-based treatment protocols while ensuring appropriate escalation of care when necessary [43].

Pharmacists play a critical role in antimicrobial stewardship (AMS) programs, serving as key stakeholders in optimizing antimicrobial use and combating resistance. In hospital settings, pharmacists perform various stewardship activities including prospective audits, formulary management, de-escalation, guideline development, and education. Pharmacist-led interventions significantly improve antibiotic prescribing, reduce unnecessary antibiotic use, and enhance patient outcomes [44]. In sepsis management, pharmacists contribute to multidisciplinary teams by expediting antibiotic delivery, ensuring appropriate selection, and improving time to administration. Studies demonstrate that pharmacist involvement in sepsis response teams reduces time to antibiotic administration from hours to minutes and increases the proportion of patients receiving appropriate antibiotics within the first hour [41,45]. Pharmacists also assist with fluid management and vasopressor facilitation for severe cases [41]. Future potential roles include expanded community-based AMS initiatives, leadership in transitions of care, implementation of biomarker-guided therapy, and development of technology-driven solutions for surveillance and patient engagement [46]. Standardized AMS training and pharmacy curricula modification are needed to enhance pharmacists’ capabilities in these critical areas [47]. Interprofessional collaboration significantly improves sepsis guideline adherence and patient outcomes through coordinated multidisciplinary approaches [48,49,50]. Studies demonstrate that collaborative sepsis teams increase bundle compliance from 25% to 62% while reducing mortality ratios from 1.14 to 0.73 [49]. Implementation of interdisciplinary code sepsis teams achieves notable improvements in fluid resuscitation compliance, lactate collection timing, and mortality reduction from 12% to 5% [50]. Multidisciplinary educational initiatives enhance provider knowledge and foster interprofessional understanding of roles. Virtual telesimulation programs facilitate application of mental models and communication strategies in clinical practice [33]. Comprehensive sepsis programs incorporating multidisciplinary committees, sepsis champions, and real-time audits demonstrate sustained improvements in care processes [49].

## Limitations

Despite the comprehensive insights generated, this study has several limitations that should be acknowledged. First, the cross-sectional design only provides a snapshot of the participants’ knowledge, attitudes, and practices (KAP) at a single point in time. It does not allow for causal inferences or assessment of changes over time. Second, the use of a self-administered questionnaire may introduce social desirability bias, where participants might overreport positive behaviours or attitudes related to sepsis management. Additionally, the online mode of data collection may have allowed participants to look up answers, particularly in the knowledge section, potentially inflating the accuracy of responses. The sample was drawn from a limited number of healthcare facilities, which may affect the generalizability of findings to the broader population of healthcare professionals in other regions or countries. Furthermore, while the study captured a range of professional groups, the number of physicians was relatively small compared to nurses, possibly limiting comparative strength in subgroup analyses. Lastly, qualitative insights from interviews or focus groups were not incorporated, which could have provided deeper context to the observed quantitative findings. Future studies should consider longitudinal and mixed-methods designs to address these gaps and validate the current findings more robustly.

## Conclusions

The knowledge in sepsis and septic shock management appeared to be generally moderate to high amongst healthcare professionals. However, there is a gap of knowledge in few of the critical aspects, while the various basic fundamental aspects such as early recognition and administration of antibiotics were well understood. It is notable that less than half of the respondents correctly identified the role that the qSOFA score plays in screening for the risk of sepsis, the current screening tool used to identify risk for sepsis. In addition, uncertainty concerning the measurement of lactate and its use for monitoring tissue perfusion where sepsis may be present also indicates a need for greater importance to be placed on their biochemical markers.

Although the majority of respondents correctly responded to the need for inotrope, vasopressor support and fluid resuscitation, they were uncertain about whether inotropes should be used in the setting where vasopressors and fluids fail to achieve adequate circulation. Furthermore, the confusion in the metabolic management targets, e.g., of glucose and hemoglobin levels, also indicates the knowledge gap in the advanced sepsis management. These deficiencies imply that even though healthcare professionals have a clear understanding of some core principles used for these types of decisions in septic critically ill patients, they need further and more specialized training to become proficient in making such complex decisions.

The study also showed that participants were overwhelmingly positive on sepsis management attitudes. The vast majority of people understood the significance of early recognition and treatment, stressing the role of rapid intervention in enhancing patient outcomes. Confidence varied in identifying sepsis, with a considerable number indicating uncertainty, suggesting the requirement for further training and clinical exposure in order to promote self-efficacy. Furthermore, while many respondents stated that sepsis guidelines are clear and helpful, a significant number were neutral or found their implementation difficult, suggesting some barriers, such as difficulty in obtaining the guidelines, not having adequate institutional support, or workload of the clinical team.

The results on the clinical practice in sepsis management were mixed. It was encouraging to see a majority of respondents practice key evidence-based practice, such as early administration of antibiotics, collection of blood cultures, identification of the source and frequent patient re evaluation. Nevertheless, in several areas of practice, gaps were observed such as sepsis screening tool use (e.g., qSOFA), routine assessment of fluid responsiveness using ultrasound, and the use of corticosteroids and blood products under certain circumstances. Furthermore, a major proportion of participants did not monitor coagulation parameters consistently, indicative of not being able to identify Disseminated Intravascular Coagulation (DIC), a reported sepsis complication, promptly.

## Data Availability

For Study Protocols: No datasets were generated or analysed during the current study. All relevant data from this study will be made available upon study completion.

## Acknowledgments

Not applicable.

